# Strategies to improve uptake of adolescent family planning and post-abortion care services in Northern Uganda: a qualitative descriptive study

**DOI:** 10.64898/2026.09.24.26363969

**Authors:** Maxson Kenneth Anyolitho, Bernard Omech, Eustes Kigongo, Edmonton Acheka, Judith Abal Akello, Morris Chris Ongom, Samson Udho, Murara Odette, Walter Acup

## Abstract

**Introduction:** Adolescents in low- and middle-income countries continue to face barriers to family planning (FP) and post-abortion care (PAC), including stigma, limited confidentiality, inadequate information, and health-system constraints. This study explored adolescents’ and key stakeholders’ perspectives on strategies to improve uptake of adolescent FP and PAC services in Lira District, Northern Uganda.

**Methods and Materials:** An exploratory qualitative descriptive study was conducted using 13 focus group discussions (FGDs) with 65 adolescent citizen scientists aged 10-19 years and 13 key informant interviews (KIIs) with stakeholders involved in adolescent sexual and reproductive health and PAC. Data were collected using semi-structured guides, audio-recorded with consent, and supplemented by field notes, then analyzed thematically using Braun and Clarke’s six-phase framework.

**Results:** Five interconnected themes emerged, including strengthening school, community, and faith-based SRH education; strengthening the health system and service delivery; policy, legal, and institutional clarity; community engagement and trust building; and integration of FP and PAC. Participants prioritized accessible and accurate SRH education, supportive community engagement, confidential and respectful care, and reliable commodities, clear operational guidance, coordinated referrals, and integrated services. The findings indicate that uptake is shaped by interacting with individual, social, health-system, and policy conditions rather than knowledge alone. A participatory, multilevel approach that connects community-generated knowledge with adolescent-responsive health-system strengthening may offer a feasible pathway for improving FP and PAC uptake in Northern Uganda.

**Conclusion:** Adolescents and key stakeholders identified interconnected community, health-system, policy, and service-delivery strategies for improving uptake of FP and PAC services. Priority actions included accessible SRH education, supportive community engagement, confidential and respectful care, reliable commodities, clear operational and legal guidance, and integrated FP and PAC delivery.

## 1. Introduction

Adolescents in low- and middle-income countries face substantial risks of unintended pregnancy and unsafe abortion, with barriers spanning information, social norms, autonomy, stigma, and health-system access (1–4). Adolescents also report difficulties obtaining confidential, respectful and acceptable services(5) .Improving uptake therefore requires more than increasing knowledge; it requires attention to the social and service environments in which adolescents make reproductive-health decisions.

In Uganda, adolescent contraceptive use remains low and unmet need high, while continuity between post-abortion care (PAC) and contraception remains incomplete (6). Negative provider attitudes, privacy concerns, misinformation, stigma and uncertainty about access can discourage adolescents from seeking care (7) .Strengthening the connection between accurate information, confidential services and voluntary contraceptive choice is therefore important for preventing unintended pregnancy and supporting adolescents who require PAC.

Northern Uganda, including the Lango sub-region, faces additional constraints related to poverty, geography, health-system capacity and sociocultural norms. Adolescents may rely on informal sources of information and care, while parental and caregiver influence can shape health-seeking behaviour (8). Yet adolescents and local stakeholders remain under-represented in the design of context-specific responses.

In Lira District, these challenges occur despite the availability of health facilities providing reproductive-health services. Existing evidence points to gaps in adolescent-friendly care, confidentiality, continuity and stakeholder responsiveness. Many programmes have also relied on top-down approaches that insufficiently incorporate young people and community actors into decision-making (9–12).

Participatory approaches can improve the fit between services and community realities by involving adolescents, providers, teachers and community actors in identifying barriers and developing feasible solutions (13, 14). However, limited qualitative evidence describes the strategies that local stakeholders themselves consider feasible for improving adolescent FP and PAC uptake.

The Comprehensive Adolescent Family Planning and Post-Abortion Care (CAFFP-PAC) initiative was established in Lira District using citizen science and community-led engagement to position adolescents and community actors as co-creators of locally relevant solutions (1, 15, 16). Building on this initiative, the study explored adolescents’ and key stakeholders’ perspectives on strategies for improving uptake of adolescent FP and PAC services in Lira District, Northern Uganda.

Lira University and its partners launched the Comprehensive Adolescent Family Planning and Post-Abortion Care (CAFFP-PAC) initiative in Lira District. Using citizen science and community-led engagement, the initiative positions adolescents and community actors as co-creators of locally relevant ASRH solutions (1, 15, 16). The project conducted baseline assessments of knowledge, attitudes, practices, and barriers and facilitators to the uptake of AFFP and PAC services through surveys, FGDs, and KIIs, with findings to be reported in another manuscript. Building on the identified barriers, we explored adolescents’ and key stakeholders’ perspectives on strategies for improving uptake of adolescent family planning and post-abortion care services in Lira District, Northern Uganda.

## 2. Methods and materials

### 2.1 Study design

This exploratory qualitative descriptive study examined adolescents’ and key stakeholders’ perspectives on strategies for improving uptake of adolescent family planning (FP) and post-abortion care (PAC) services in Lira District, Northern Uganda. Data were generated through focus group discussions (FGDs) with adolescent citizen scientists aged 10-19 years and key informant interviews (KIIs) with purposively selected stakeholders involved in adolescent sexual and reproductive health (ASRH) and PAC. The study was embedded within the CAFFP-PAC initiative, which used a participatory, community-led approach.

### 2.2 Study site and setting

The study was conducted in Lira District in the Lango sub-region of Northern Uganda. Lira was selected because of its adolescent reproductive-health needs and challenges affecting access to confidential, high-quality and adolescent-responsive services. The setting includes rural and urban communities and primary healthcare facilities serving adolescents (17) (1, 18, 19).

### 2.3 Study population, sample size and sampling

The study included adolescents aged 10-19 years participating as citizen scientists and stakeholders involved in ASRH and PAC, including health workers and community actors. The final dataset comprised 26 qualitative sessions involving 78 individuals: 13 FGDs with 65 adolescents and 13 KIIs with 13 stakeholders. Participants were purposively selected to obtain diversity in age, gender, roles and involvement in adolescent health and service delivery. Sample size was determined iteratively using the study team’s assessment of saturation.

### 2.4 Data collection

Thirteen FGDs with 65 adolescent citizen scientists explored experiences, social norms, perceived barriers and suggested strategies. Thirteen KIIs with stakeholders explored service delivery, community influences and institutional strategies. Semi-structured, pre-tested guides were used. Sessions were conducted in private or neutral locations, in English or Lango according to participant preference, and were audio-recorded with consent and supplemented by field notes.

### 2.5 Research team and reflexivity

Four research assistants (two female and two male), fluent in English and Lango, facilitated data collection. They were bachelor’s-level in public health (Ocan Dennish-OD) and social science (Akello Christin-AC, Okello Jimmy-OJ and Grace Dorrothy Atino-GDA) graduates with at least five years’ qualitative data-collection experience and received two days of study-specific training. They had no prior relationships with participants. For KIIs, each assistant facilitated and took notes; for FGDs, assistants worked in facilitator/note-taker pairs and rotated roles across sessions. Before each session, participants were informed about the research team’s role, the study purpose and the voluntary nature of participation. The team-maintained separation between participants’ accounts and researchers’ interpretations through team review of the coding framework, triangulation and an audit trail.

The research team’s roles were primarily facilitation, note-taking and analysis. No prior relationships with participants were reported, and private or neutral settings were used to support candid discussion.

### 2.6 Study Procedure

Following ethical approval and community entry, district and sub-county leaders were engaged to facilitate access to the study. Potential participants were purposively identified and contacted, and sessions were scheduled for convenience and privacy.

Before each session, written informed consent and assent were obtained from participants. Research assistants explained the study’s purpose, scope, and voluntary nature. For adolescent citizen scientists aged 13-17 years, written assent was obtained as part of the informed consent process, and written parental consent was also obtained from their parents/caregivers. For adolescents aged 18 and 19 years, and for local leaders, written informed consent was obtained. All consent forms were translated into the local language (leb lango) to facilitate interpretation and understanding by participants and parents/caregivers with limited or no education. No non-participants were present during the sessions. Sessions lasted approximately 40–90 minutes. Of 78 adolescents mobilized for FGDs, 65 participated; 13 did not participate because they were not at home or were occupied with gardening and other tasks. No repeat interviews or FGDs were conducted. Data collection occurred from June to September 2025. Potential sources of bias included selection and social-desirability bias; these were addressed through purposive sampling for diversity, trained facilitators without prior relationships with participants, private settings, use of both FGDs and KIIs, and field notes and team-based triangulation.

### 2.7 Data Analysis

Data were analyzed thematically using Braun and Clarke’s six-phase framework(20, 21). Audio recordings and field notes were transcribed verbatim and, where applicable, translated into English while retaining the meaning of participants’ accounts. No transcripts were returned to participants. Before coding, all transcripts were uploaded to NVivo software version 15. As the analysis was done by the research team, no participant checked the transcripts or the codes except for the research team itself. The analysis involved familiarization with the data, generation of initial codes, searching for candidate themes, reviewing and refining themes, defining and naming themes, and producing the final analytic account. Coding focused on participants’ experiences and proposed strategies for improving adolescent FP and PAC uptake. Data from FGDs and KIIs were compared and triangulated to identify convergent and divergent perspectives across participant groups. To enhance analytic rigour, the coding framework was developed by the corresponding author and reviewed and refined by three other members of the research team, with discrepancies discussed and resolved through consensus. Themes were supported by illustrative participant quotations. An audit trail and triangulation of data sources, transcripts, recordings, and field notes were used to enhance credibility, dependability, and confirmability.

#### 2.7.1 Data adequacy and analytic decision-making

Data collection and analysis were iterative, with questioning refined as findings emerged. The final sample was considered adequate by the research team based on saturation. No formal information-adequacy framework was applied.

#### 2.7.2 Reporting guideline

Reporting was cross-checked against the Consolidated Criteria for Reporting Qualitative Research (COREQ), the primary framework for this qualitative study. STROBE was used only as a supplementary transparency check because it was developed for observational epidemiological studies.

### 2.8 Ethics Statement

The study protocol was reviewed and approved by the Lira University Research Ethics Committee (LUREC-2024-309) and registered with the Uganda National Council for Science and Technology (UNCST-HS6030ES). The study was conducted in accordance with the principles of the Declaration of Helsinki. The ethics committee approved written informed consent. Before participation, caregivers were informed of the study purpose, participants’ rights to withdraw without penalty, and the use of anonymized quotations. Participant confidentiality and secure data storage were maintained throughout the study.

## 3. Results

### 3.1 Participant characteristics

The study included 78 individuals across 26 qualitative sessions: 65 adolescents in 13 FGDs and 13 stakeholders in 13 KIIs. Among KII participants, 8 (61.54%) were female and 5 (38.46%) male; 9 (69.23%) were health workers, 2 (15.38%) VHTs, 1 (7.69%) peer educator and 1 (7.69%) student. Among FGD participants, 47 (72.31%) were female and 18 (27.69%) male; 55 (84.62%) were aged 15 years or older and 38 (58.46%) were students. Participant characteristics are presented in Table 1.

**Table 1:**

| Participant group / characteristic | Category | n | % |
| --- | --- | --- | --- |
| KII participants (n=13) |  |  |  |
| Gender | Female | 8 | 61.54 |
|  | Male | 5 | 38.46 |
| Age | Below 36 years | 6 | 46.15 |
|  | 36 years and above | 7 | 53.85 |
| Education | Secondary | 3 | 23.08 |
|  | Post-primary | 1 | 7.69 |
|  | Tertiary | 9 | 69.23 |
| Occupation | Health worker | 9 | 69.23 |
|  | Peer educator | 1 | 7.69 |
|  | Student | 1 | 7.69 |
|  | VHT | 2 | 15.38 |
| Religion | Anglican | 5 | 38.46 |
|  | Catholic | 2 | 15.38 |
|  | Born Again/Protestant | 6 | 46.15 |
| FGD participants (n=65) |  |  |  |
| Gender | Female | 47 | 72.31 |
|  | Male | 18 | 27.69 |
| Age | Below 15 years | 10 | 15.38 |
|  | 15 years and above | 55 | 84.62 |
| Education | Primary | 43 | 66.15 |
|  | Secondary | 22 | 33.85 |
| Marital status | Married | 19 | 29.23 |
|  | Single | 24 | 36.92 |
|  | Missing | 22 | 33.85 |
| Occupation | CHEW | 3 | 4.62 |
|  | Farmer | 11 | 16.92 |
|  | Hairdresser | 1 | 1.54 |
|  | Key population | 9 | 13.85 |
|  | Student | 38 | 58.46 |
|  | Tailoring | 1 | 1.54 |
|  | PSW | 1 | 1.54 |
|  | Secretary | 1 | 1.54 |
| Religion | Anglican | 19 | 29.23 |
|  | Catholic | 21 | 32.31 |
|  | Muslim | 1 | 1.54 |
|  | Pentecostal | 14 | 21.54 |
|  | SDA | 3 | 4.62 |
|  | Missing | 7 | 10.77 |

*Source: Data from participants, 2025*

**Fig 1.**
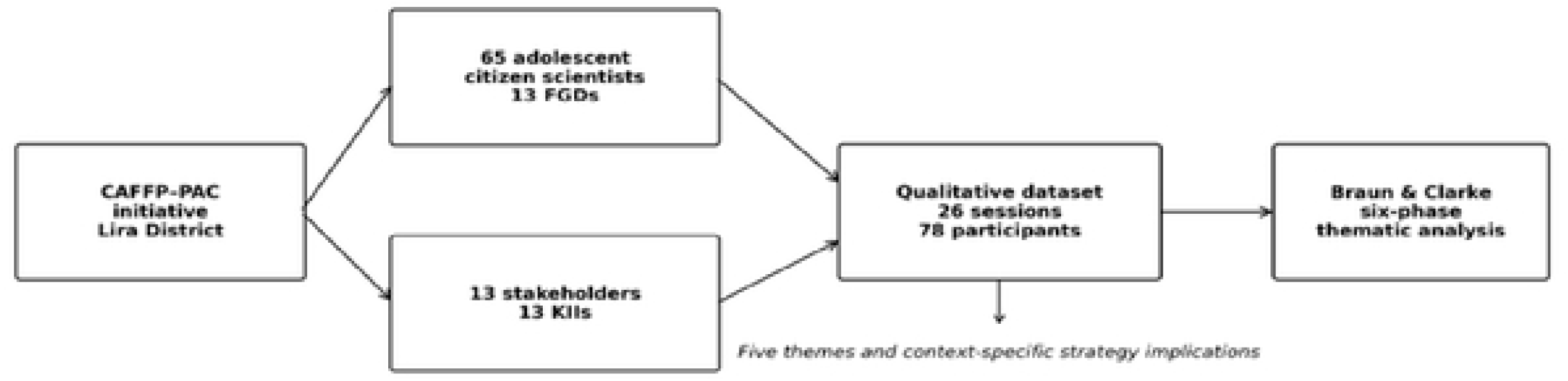
Study participant and analytic pathway. The analytic pathway from the CAFFP-PAC initiative through the two qualitative data sources to thematic synthesis is shown in Figure 1. The diagram shows the 26 qualitative sessions, 78 participants and the thematic analysis pathway. It is a descriptive analytic schematic rather than a causal model.

**Fig 2.**
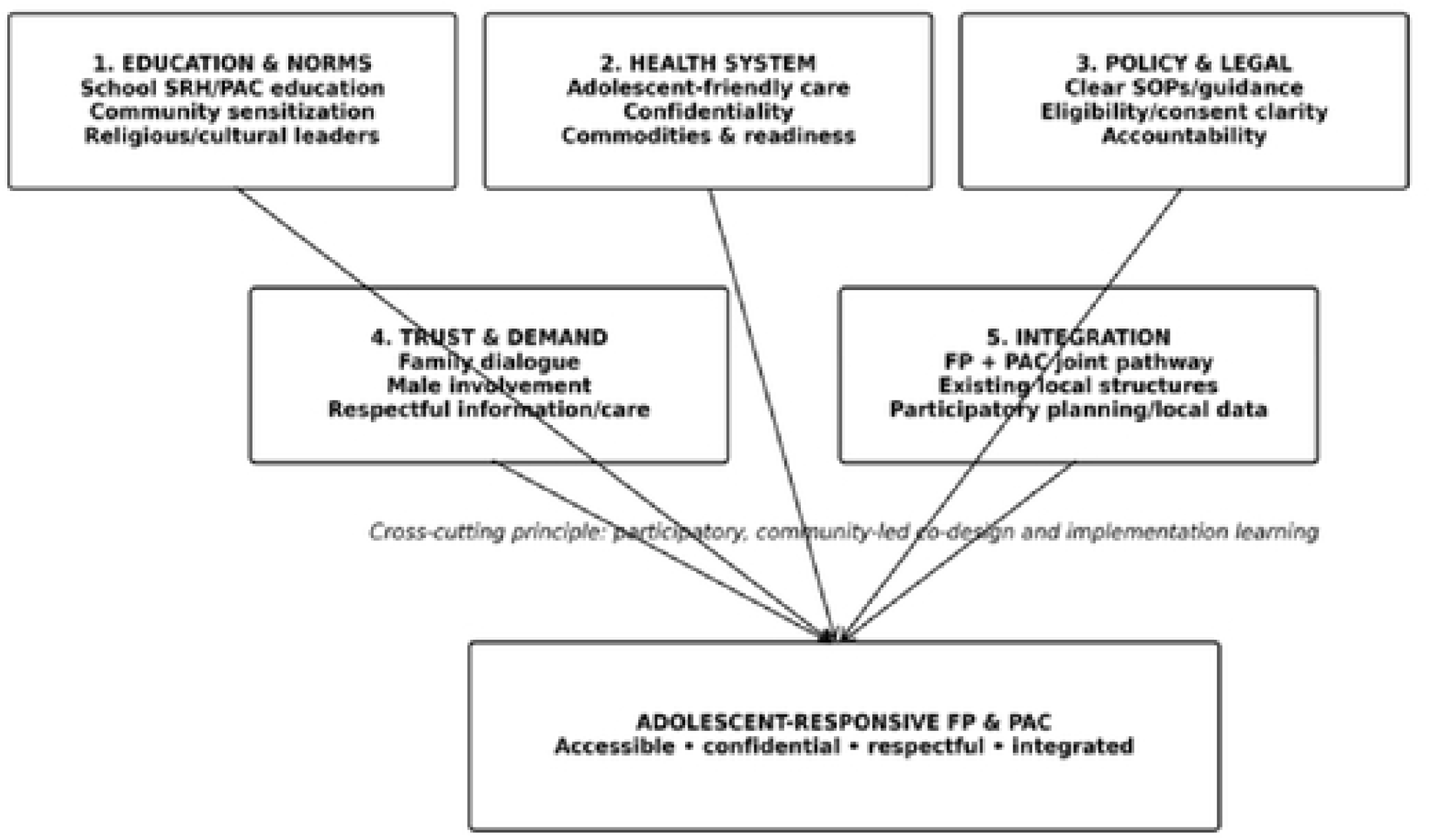
Multilevel strategy framework emerging from adolescent and stakeholder perspectives. The framework is an analytic synthesis of the five themes and is not a quantitatively tested causal model.

**Fig 3.**
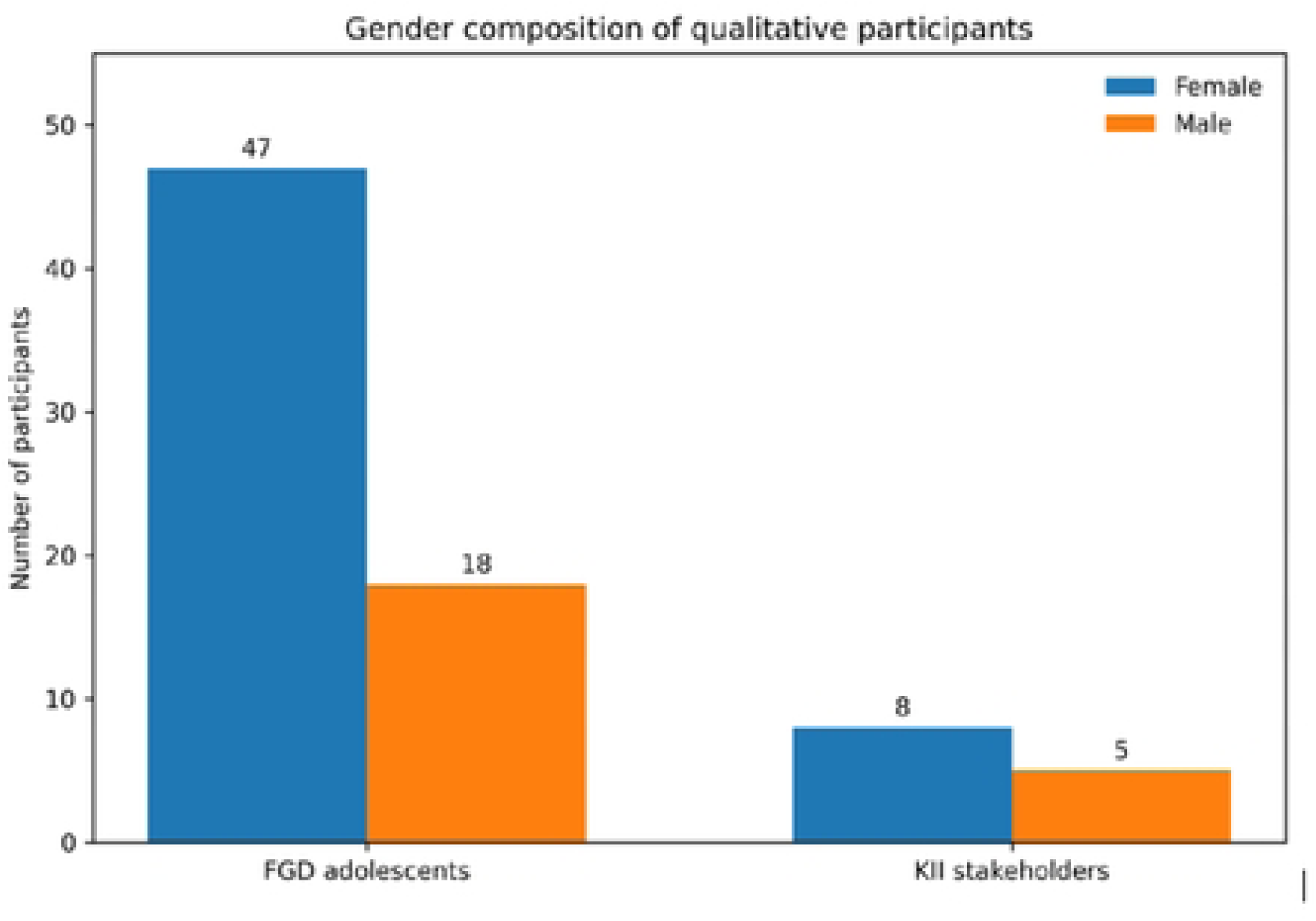
Gender composition of qualitative participants. Bars show participant counts by qualitative data source and sex/gender as reported in the descriptive dataset.

**Table 2.** Summary of themes, sub-themes and proposed strategy implications.

| Theme | Sub-themes | Proposed strategy implications |
| --- | --- | --- |
| **1. Education and norms** | School SRH/PAC education; community sensitization; religious and cultural leader engagement | Expand structured school outreach and health clubs; conduct community dialogues; engage trusted leaders in supportive messaging. |
| **2. Health system** | Adolescent-friendly care and confidentiality; commodity availability; coordinated stakeholder collaboration | Maintain private adolescent-friendly spaces; strengthen provider capacity; prevent stock-outs; improve referral and coordination. |
| **3. Policy and legal** | Clear policies/SOPs; legal clarity and protection | Disseminate actionable guidance; clarify eligibility, consent/guardian issues, and access to life-saving PAC; strengthen accountability. |
| **4. Trust and demand** | Family dialogue and male involvement: information and respectful care | Promote non-judgmental family dialogue; engage male champions; provide clear information; strengthen respectful care. |
| **5. Integration** | Coordinated service delivery; existing local structures; participatory planning and local data | Integrate FP and PAC at the point of care; leverage existing platforms; use community-generated evidence for co-design and adaptation. |

***Note:** This synthesis represents the qualitative interpretation of participant recommendations and* *is not a tested causal framework.*

### 3.2 Theme 1: Strengthening School, Community, and Faith-Based SRH education and sensitization

#### Sub-theme 1.1: School-based SRH and PAC education

Participants recommended integrating FP and PAC education within schools through regular engagement of trained health workers and structured outreach programs. They emphasized that schools provide a strategic platform for reaching adolescents early with accurate ASRH information before risky behaviors start to occur. School-based sessions were seen as an opportunity to normalize discussions on FP and PAC while addressing myths and misconceptions. Participants further suggested interactive approaches such as demonstrations, health talks, and establishing school health clubs to enhance understanding and retention of information. This integration was viewed as essential for improving awareness, informed decision-making, and timely uptake of ASRH services

> *“School head teachers should invite health workers to teach and talk to adolescents about sexual and reproductive health so that our children can learn directly from professionals.”***(FGD-Rapha-In-school 15-19, P01)**
>
> *“School outreaches with structured health education sessions, including practical demonstrations, would help students better understand and apply what is taught.”***(KII Master Coach-Aromo HCIII)**

#### Sub-theme 1.2: Community sensitization/engagement and stigma reduction

Participants emphasized the importance of regular community dialogues as a way to normalize help-seeking behaviours among adolescents for FP and PAC services. They noted that open discussions within communities can help break the silence around sensitive SRH issues. Such dialogues were seen as critical in reducing stigma and misconceptions associated with contraceptive use and post-abortion care. Participants further highlighted that involving respected community members in these conversations could increase acceptance and trust. Participants also believed that community engagement would create a more supportive environment in which adolescents could access care without fear or judgment.

> *“Community leaders should organize meetings and inform the community about adolescent health services so that people are well informed.”***(FGD-In-school 15-19, Rapha)**
>
> *“Community meetings with health workers help us a lot because we learn many things about how these services work for our young children and encourage them to stop fearing to go for FP and PAC services.”***(FGD-In-school 15-19, Rapha)**

#### Sub-theme 1.3: Engagement of religious and cultural leaders

Religious and cultural leaders were viewed as key actors in shaping community norms and influencing attitudes toward adolescent sexual and reproductive health. Their voices were seen as powerful in either reinforcing or reducing stigma associated with FP and PAC services. Participants emphasized that when these leaders provide supportive and non-judgmental messages, community acceptance and adolescents’ willingness to seek care significantly improve.

> *“You see if pastors and elders speak positively about going to the hospital for treatment, many people in the community will be encouraged to seek care.”***(FGD-Aromo)**
>
> *“If elders and church leaders talk about hospital care in a kind and supportive way, we will not be afraid to go for FP services.”***(FGD-Aromo)**
>
> *“Cultural and religious leaders should be engaged to include PAC sensitization in their regular community and religious gatherings.”* (KII-Peer Educator, Lira District)

### 3.3 Theme 2: Health System Strengthening and Service Delivery Improvement

#### Sub-theme 2.1: Adolescent-friendly service delivery and confidentiality

Participants emphasized the need for clear and well-communicated guidelines on ASRH services to reduce confusion and fear. They highlighted confidentiality as a critical factor that would encourage adolescents to seek FP and PAC services without fear of exposure or being judged. Adolescent-friendly spaces within health facilities were also seen as essential for creating a safe and welcoming environment. Strengthening these aspects would significantly improve trust in FP and PAC services and increase uptake among adolescents.

> *“Strong policy support is needed to ensure consistent provision of adolescent-friendly services and uninterrupted availability of essential commodities.”***(KII-Master Coach-Aromo HCIII)**
>
> *“If we are clearly informed about what services are free at the different health facilities and which ones require a guardian, it would reduce fear and confusion among adolescents.”***(FGD-CS, Aromo)**

#### Sub-theme 2.2: Commodity availability and service readiness

Consistent availability of FP commodities and PAC services was seen as essential for improving access and continuity of care for adolescents. Participants emphasized that stock-outs and limited-service points significantly hinder timely utilization of ASRH services among adolescents. They noted that reliable supply systems would increase trust in public health facilities.

> *“Government should provide more services and adequate commodities at all health facilities.”***(FGD-In-school 15-19, Rapha)**

Consistently ensuring steady availability of both services and supplies was viewed as a key driver of improved adolescent uptake of FP and PAC services.

#### Sub-theme 2.3: Coordinated stakeholder collaboration

Participants emphasized the importance of strong coordination between health workers, non-governmental organizations (NGOs), and community actors to improve the delivery of adolescent SRH services. Collaboration would help reduce duplication of efforts and improve efficiency in ASRH service provision. Such coordination was seen as key in strengthening referral systems and ensuring continuity of care for adolescents. Additionally, participants believed that joint planning and shared implementation would enhance resource utilization and service coverage.

> *“Enhance coordination by bringing all relevant stakeholders together for joint planning and implementation and ensure that the adolescent corner are kept open and functional every day to serve young people effectively.”***(KII-Master Coach-Aromo HCIII)**
>
> *“Collaboration between government and non-governmental organizations such as RHU, GLOFORD, and DREAMS/TCI is essential in strengthening and sustaining adolescent health services.”***(KII-Midwife, Ober HCIV)**

### 3.4 Theme 3: Policy, Legal, and Institutional Strengthening

#### Sub-theme 3.1: Clear policies and operational guidelines

Stakeholders called for clear, actionable policies and well-defined standard operating procedures (SOPs) to guide the delivery of adolescent sexual and reproductive health (SRH) and post-abortion care (PAC) services. They emphasized that such guidelines would help standardize service provision across facilities and reduce inconsistencies in care. Participants also noted that clear policies would support health workers in making appropriate decisions while ensuring adolescent rights are upheld. In addition, they believed that actionable SOPs would improve accountability and service quality within health facilities. This was seen as essential for strengthening adolescent-friendly service delivery systems.

> *“Implement clear guidelines and reforms for adolescent health services to ensure standardized and youth-responsive care.” ………“Ensure consistent availability of essential supplies to avoid interruptions in service delivery for adolescents.” ………“Establish and maintain dedicated adolescent corners within health facilities to provide safe, private, and friendly spaces for care.”* **(KII-Master Coach-Aromo HCIII)**

#### Sub-theme 3.2: Legal clarity and protection for adolescents

Participants highlighted the need for clear legal guidance on ASRH and PAC services to reduce fear among both providers and clients. They noted that legal clarity would improve timely access to care by minimizing delays caused by uncertainty and stigma.

> *“Clarify legal access questions for under-18s, prioritize life-saving PAC.”***(KII-Master Coach-Aromo HCIII)**
>
> *“If they tell us what is free and what needs a guardian, it will reduce fear.”***(FGD-CS, Aromo)**

### 3.5 Theme 4: Community Engagement, Trust Building, and Demand Creation

#### Sub-theme 4.1: Family, caregiver, and male involvement

Participants emphasised the importance of strengthening communication within families to improve understanding and support for ASRH needs. They noted that open dialogue between parents and adolescents can reduce misinformation and stigma around FP and PAC services. Engaging male champions was also seen as critical in promoting shared responsibility and positive attitudes toward adolescent health. These approaches were viewed as key to creating a supportive environment for informed decision-making and service uptake.

> *“Engaging both parents and male champions in adolescent health discussions helps improve communication and strengthens support for ASRH services at family level.”***(FGD-Aromo)**

This finding highlights the perceived importance of family dialogue and male involvement in creating a supportive environment for adolescent ASRH.

#### Sub-theme 4.2: Trust-building through information and respectful care

Confidence in ASRH services was strongly linked to the assurance of privacy within health facilities and service providers. Participants emphasized that clear and understandable information about available services builds trust and encourages service use among adolescents. Respectful and non-judgmental treatment by health workers was viewed as essential for sustaining adolescents’ confidence in seeking care.

> *“Community meetings with health workers help us a lot because we learn many things from them, and they also help us understand SRH issues better and correct the wrong information we had, so through these meetings we stop fearing to go for services like FP and PAC.”***(FGD-In-school 15-19, Rapha)**

### 3.6 Theme 5: Integration Strategies for CAFFP-PAC in Primary Healthcare

#### Sub-theme 5.1: Coordinated and accessible service delivery

Stakeholders emphasized the importance of integrating FP and PAC services at the same point of care to improve efficiency and continuity of services. They noted that providing both services together would reduce missed opportunities for counseling and method uptake.

Integration was also seen to minimize multiple facility visits, which often contributes to dropout among adolescents.

> *“Integration means family planning and post-abortion care are available at the same place and time, reducing missed opportunities. If a girl comes for care and must return another day for counseling, she may never come back.”***(KII-Health worker)**

#### Sub-theme 5.2: Leveraging existing local structures

Participants recommended strengthening existing school clubs and community structures rather than creating parallel systems for ASRH programming. They emphasized that school health clubs and community groups that already exist can be leveraged effectively for FP and PAC sensitization. Integrating SRH activities into these platforms was seen as a cost-effective and sustainable approach.

> *“We already have school health clubs and community groups. What’s needed is to bring them together with health centers, and they start sensitizing the public to FP and PAC services and where to get them. They only need to be provided with the right information.”***(CBO representative)**

#### Sub-theme 5.3: Participatory planning and use of local data

Participants highly valued community engagement as it helped to reveal real-life barriers affecting adolescents’ access to FP and PAC services. They noted that direct interaction with community members provided a deeper understanding of socio-cultural and structural challenges. This engagement was also seen as essential for co-creating practical and context-specific solutions that reflect local realities.

> *“The inception meeting helped uncover the social, cultural, and structural barriers that need to be addressed together. Also, without listening to us, they would never have known how hard it is to reach the clinic from our village.”***(KII-District official)**

## 4. Discussion

This study identified five interconnected strategy areas for improving adolescent FP and PAC uptake in Lira: SRH education and norm change; health-system strengthening; policy and legal clarity; community engagement and trust; and FP-PAC integration. Together, the findings indicate that uptake is not simply a matter of knowledge or motivation. It is shaped by whether adolescents can act on information within supportive social environments and reach services that are confidential, respectful, available and easy to navigate. This multilevel interpretation is consistent with recent evidence from sub-Saharan Africa on individual, sociocultural, health-system and policy barriers and on adolescents’ preferences for confidential, convenient and respectful care (22–24).

### 4.1 Education, knowledge, and the social production of reproductive-health behaviour

Participants’ emphasis on school-based education, community sensitization and religious or cultural leaders indicates that information operates within a wider social environment. Adolescents may know FP or PAC exist but still be uncertain where to obtain care, whether confidentiality will be protected, or whether using services will attract judgments. Recent evidence similarly identifies misinformation, fear of disclosure, parental disapproval, stigma and restrictive norms as interacting barriers (23, 25). Education should therefore also make service pathways visible and strengthen adolescents’ confidence to seek care.

Schools offer a structured platform for age-appropriate information and referral. The 2025 WHO guideline places education, contraception and supportive environments within a broader package for preventing early pregnancy (26, 27). However, education is unlikely to increase FP-PAC uptake when services remain difficult to access. It should be linked to clear information about where services are available, what adolescents can expect and how confidentiality is protected.

Religious and cultural leaders are part of the normative environment rather than merely communication channels. Their messages may reduce shame and normalise appropriate care or reinforce silence when they are moralising. Engagement should therefore be assessed by whether it produces accurate, rights-respecting and non-stigmatising messages. This aligns with WHO standards that place adolescent empowerment, family/community engagement and welcoming services alongside competent care(26).

### 4.2 Health-system readiness: from “youth-friendly” labels to experienced quality of care

The second major finding concerns what adolescents experience when they reach a health facility. Participants prioritised confidentiality, adolescent-friendly spaces, respectful providers, trained staff, commodity availability and coordinated referrals. Recent evidence shows that judgmental provider attitudes, inadequate privacy, poor accessibility and contextual constraints remain common across sub-Saharan Africa (23, 24, 28).

“Adolescent-friendly” should therefore be understood as an experienced quality-of-care construct, not simply the presence of a youth corner. A facility can have a designated space yet remain inaccessible if adolescents fear being recognised, judged or turned away because a method is unavailable. This is particularly important for PAC, where disclosure and stigma may be heightened. WHO standards similarly emphasise accessibility, acceptability, equity, privacy, confidentiality and respectful care (26).

Commodity availability links supply-side readiness to future demand. An unsuccessful service encounter can delay contraceptive initiation and undermine trust. Evidence from Southwestern Uganda found gaps in staffing, equipment and adolescent-service capacity, alongside substantial training needs in adolescent-friendly and PAC care (29). Training should therefore combine clinical competencies with communication, confidentiality, informed choice and non-discrimination. The service-strengthening package emerging from this study is consequently broader than infrastructure: competent providers, private consultation, reliable commodities, functional referrals, supportive supervision and routine monitoring of adolescents’ experience of care.

### 4.3 Policy and legal clarity: the implementation gap matters as much as the policy itself

Participants’ call for clear policies, SOPs and legal guidance shows how policy ambiguity can become an everyday service-delivery barrier. Providers encounter policy through decisions at registration, consultation and referral points. Uncertainty about eligibility, confidentiality, consent or independent access can result in delay or referral elsewhere. Recent evidence and WHO guidance similarly identify policy restrictions and unclear procedures as barriers to adolescent contraception and responsive care (23, 26).

The implication is not simply to develop new policies but to operationalise existing guidance through accessible SOPs, provider orientation, decision-support tools and supportive supervision. The legal concerns raised by participants particularly regarding access for adolescents younger than 18 years, should be interpreted as perceived uncertainty rather than a legal interpretation of Uganda’s abortion framework. For PAC, the practical priority is to minimise avoidable delays in medically indicated care while ensuring that providers have clear, lawful guidance.

### 4.4 Family, gender, and community trust: support without undermining adolescent autonomy

The fourth theme highlights a tension in adolescent SRH programming: parents, caregivers, partners and community actors can provide support, but the same relationships can also generate surveillance, stigma or control. Participant recommendations for family dialogue and male involvement should strengthen adolescents’ support environments without transferring decision-making authority away from them. This is consistent with evidence that social support, information, accessibility and youth involvement are interconnected components of adolescent SRH needs (23, 24).

Male involvement should promote shared responsibility, communication and respect rather than control over contraceptive decisions. Trust emerged as a bridge between community engagement and service use: adolescents linked trust to understandable information, respectful treatment and protection of privacy. Community engagement should therefore be judged by whether it reduces stigma, improves accurate information and creates safe routes to care. Adolescents should also have opportunities to provide feedback on services.

### 4.5 Integration of FP and PAC: reducing missed opportunities across the continuum of care

Integrating FP and PAC at the point of care was one of the clearest service-delivery recommendations. Participants viewed integration to reduce repeat visits, missed counselling opportunities and dropout. Evidence supports this pathway: post-abortion FP counselling is associated with contraceptive use, while education, prior FP use and partner support also influence uptake (30, 31).

Integration should mean more than co-location. It requires coordination of providers, counselling, commodities and referrals so that an adolescent can move through PAC and voluntary, informed contraceptive care without unnecessary additional visits. Participants also preferred using existing school and community structures rather than creating parallel systems. For CAFFP-PAC, integration is therefore both a service-delivery strategy and an implementation principle linking community information, facility care and follow-up. Participatory evidence from sub-Saharan Africa remains limited, reinforcing the need to evaluate whether adolescent involvement improves service outcomes (32).

### 4.6 The central contribution: moving from isolated barriers to a participatory, multilevel theory of uptake

The principal contribution of the study is the way the five themes fit together. Information, social norms, trust, provider behaviour, commodity availability, policy implementation and service organisation can reinforce one another. Misinformation may increase reliance on informal sources; stigma may discourage disclosure; provider judgement may confirm community fears; and stock-outs may undermine trust after an adolescent decides to seek care. Conversely, accurate information, supportive relationships, respectful providers, reliable commodities and integrated services can create mutually reinforcing conditions for uptake.

This interpretation aligns with WHO’s adolescent-health standards, which combine empowerment and participation, rights-based care, competent providers, family/community engagement and quality improvement (33). It also aligns with participatory evidence suggesting that adolescents can contribute to intervention design while highlighting the limited evidence on whether participation improves health-system outcomes (32). The value of citizen science in CAFFP-PAC should therefore be assessed by whether adolescent-generated knowledge improves service experience, uptake, continuity and equity.

The citizen-science orientation adds an implementation-learning dimension by positioning adolescents as contributors to knowledge about barriers and feasible solutions. This can reveal practical constraints that routine facility statistics may miss, including difficulties reaching services, fear of recognition and local meanings attached to contraception. At the same time, the sample was weighted toward females and older adolescents. Future implementation should monitor differences by age, sex/gender, schooling status, location and other locally relevant characteristics so that participatory programming does not reproduce existing exclusions (22, 33).

### 4.7 Implications for implementation and policy

The findings suggest five implementation priorities. First, link school-based SRH education to clear referral pathways. Second, institutionalise confidentiality, respectful communication, provider competency, reliable FP/PAC commodities and functional referrals. Third, translate policy into concise SOPs, provider orientation and supportive supervision. Fourth, engage adolescents, parents, caregivers, male champions and community leaders while protecting adolescent autonomy and avoiding stigma. Fifth, organise FP and PAC as a coordinated continuum, with voluntary contraceptive counselling and provision during PAC where appropriate.

These priorities are consistent with current WHO guidance and evidence from Uganda on gaps in staffing, training, adolescent-specific services and PAC readiness (26, 29, 33). Because this study is qualitative and exploratory, the strategies should be evaluated in subsequent implementation phases using indicators such as adolescent satisfaction, confidentiality experience, FP initiation, PAC counselling, referral completion and continuity of use.

### 4.8 Limitations

This qualitative study provided in-depth perspectives from adolescents and stakeholders, with triangulation across 13 FGDs and 13 KIIs. The findings are context-specific and are not statistically generalisable. Because FP, PAC and adolescent sexuality are sensitive topics, social-desirability and disclosure biases may have influenced responses despite private settings and trained facilitators. The sample also contained more female and older adolescents, which may limit representation of younger adolescents and boys. Some participant characteristics had missing values, particularly marital status and religion.

The study was designed for contextual understanding rather than prevalence estimation or causal inference. Future implementation should test whether the proposed strategies improve measurable outcomes and whether effects differ across age, gender, schooling status, and other relevant groups.

## 5. Conclusion

Uptake of adolescent FP and PAC services in Lira District is shaped by interacting social, community, and health-system and policy factors. Participants prioritised accessible SRH education, supportive but non-coercive family and community engagement, confidential and respectful care, reliable commodities, clear operational guidance, coordinated referrals, and functional integration of FP and PAC. These findings support a participatory, multilevel approach that links community-generated knowledge with health-system strengthening and adolescent-responsive service delivery. Integrating FP and PAC services and strengthening existing communities and institutional structures were viewed as important strategies for reducing barriers and improving continuity of care. A participatory, multi-level approach that incorporates adolescents’ perspectives and engages relevant stakeholders is therefore critical for developing context-specific and sustainable strategies to improve adolescent FP and PAC uptake in Northern Uganda.

## 6. Declarations

## 6.1 Acknowledgments

We sincerely acknowledge the adolescents who participated in the focus group discussions and the key informants who contributed their perspectives, as well as officials from Lira University, Lira City and District Local Government, community-based organizations, teachers, youth mentors (master coaches), health workers, and community leaders who supported the study.

## 6.2 Author contributions

MKA, BO, EK, AE, JAA, MCO, SU, OM, and WA contributed to the study. BO and MKA conceptualized and designed the study. WA, EK, and AE contributed to data acquisition, analysis and interpretation. MKA and WA drafted the manuscript. SU and OM critically revised the manuscript. All authors reviewed and approved the final manuscript and agreed to its submission.

## 6.4 Data availability

The data are available from the corresponding author upon reasonable request.

